# Harmonisation of the 0-10 Numerical Rating Scale for pain intensity and the pain domain of the Western Ontario and McMaster Universities Osteoarthritis Index

**DOI:** 10.1101/2025.09.26.25336707

**Authors:** Jens Laigaard, Saber Muthanna Saber Aljuboori, Søren Overgaard, Karl Bang Christensen

## Abstract

**Background:** Analgesic efficacy is often evaluated with patient-reported outcome measures (PROMs). However, many different PROMs are used, which poses a problem when results are pooled in meta-analyses. Two commonly used PROMs used for evaluating pain intensity are the 0-10 numerical rating scale (NRS) and the pain domain of the Western Ontario and McMaster Universities Osteoarthritis Index (WOMAC).

**Objective:** In this study, we aim to evaluate if the NRS and the WOMAC pain domain can be harmonised and, if possible, to report a conversion table.

**Methods:** The study is based on a large dataset of 12-to 18-month pain outcomes after primary total hip arthroplasty (THA), total knee arthroplasty (TKA), or unicompartmental knee arthroplasty (UKA) for osteoarthritis (ClinicalTrials.gov identifiers NCT05845177 and NCT05900791). We will apply multiple imputation to impute missing WOMAC pain domain responses.

We will assess the distribution of scores and evaluate if NRS and the WOMAC pain domain have sufficient correlation and a monotonous relationship. We will also assess the NRS and WOMAC pain domain relationship within important subgroups, such as age, sex, and type of surgery. If deemed appropriate, we will perform equipercentile linking to create a conversion table from WOMAC pain domain sum scores to NRS scores and vice versa. We will assess the relationship between predicted and actual values with root mean squared errors (RMSEs) and mean absolute errors (MAEs), concordance correlation coefficients (CCCs) and intraclass correlation coefficients (ICC), Bland-Altman plots, residual plots, and calibration plots.

**Perspective:** The results will be submitted for publication in a peer-reviewed journal. We will seek to make the reports freely available, either by open-access publication or through publication on a preprint server, e.g. www.medrxiv.org.

## Introduction

Researchers undertaking systematic reviews and meta-analyses are often faced with the option to pool results from studies that have applied different patient-reported outcome measures (PROMs). Although pooling PROM data increases the information size, reporting of standardised effect sizes impairs interpretation of the estimate. Moreover, the standardisation is calculated as the difference in means between trial arms divided by the standard deviation (SD). The standardisation therefore relies on assumptions such as similar variance between study populations (i.e., that the differences in SDs reflect only differences in the PROMs), PROM validity, and that the included PROMs measure the same underlying construct.(20,21) Researchers may therefore choose to meta-analyse PROMs separately,(22) resulting in severely reduced information sizes, lower strength of evidence, and ultimately: delayed knowledge on treatment effects.(11)

In pain research, both the 0-10 Numeric Rating Scale (NRS) and the 5-item Western Ontario and McMaster Universities Osteoarthritis Index (WOMAC) pain domain are very commonly applied measures of pain.(3,12,13,23) The NRS, similar to the Visual Analogue Scale (VAS), measures pain intensity on a scale from 0 (“No pain”) to 10 (“Worst imaginable pain”), although varying scales and anchors are used.(24) In studies investigating chronic pain using the NRS, patients are typically asked to rate their average pain during the last 24 hours, or during the last week.(2,3) The WOMAC pain domain also measures pain intensity, but stratified by five different activities and rated within the last week.(4) However, in both instruments, it is uncertain if only pain intensity is measured, precluding the pain’s interference with activities, mood, social relations etc.(25) In studies assessing hip or knee osteoarthritis and arthroplasty, WOMAC is typically recommended because it is developed specifically for that population.(3) However, it is also recommended that the NRS is reported alongside the WOMAC to allow for comparison with other studies. This may be redundant if the two instruments can be harmonised soundly. Harmonisation will also facilitate meta-analyses of studies that used either instrument, thereby reducing research waste.

In this study, we aim to evaluate if the NRS and the WOMAC pain domain can be harmonised and, if possible, to report a conversion table.

### Methods Data source

In 2023, the author group undertook two surveys of measures of pain and satisfaction 12-18 months after primary total hip arthroplasty (THA), total knee arthroplasty (TKA) or unicompartmental knee arthroplasty (UKA). The surveys were registered at ClinicalTrials.gov (identifiers NCT05845177 and NCT05900791) and listed at the Capitol Region of Denmark’s research (identifiers P-2022-933 and P-2023-4). The study populations comprised 1148 UKA patients (870 respondents), 3086 TKA patients (2172 respondents), and 2777 THA patients (2031 respondents). The median time from surgery to response was 17 months (interquartile range (IQR) 16-18 months) for THA patients, and 13 months (IQR 12-14 months) for UKA and TKA patients. Among other outcomes, the surveys included the following:

- **The 0-10 numerical rating scale (NRS) for average intensity of postsurgical pain during the last week**.(2,3) *Please rate your pain in the operated knee by indicating the number that best describes your pain on average during the last week. 0 means ‘No pain’ and ‘10’ means ‘Worst imaginable pain’*.
- **The 5-item Western Ontario and McMaster Universities Osteoarthritis Index (WOMAC) pain domain (Likert-scale, version 3.1)**.(2) *What amount of knee pain have you experienced the last week during the following activities?* a) *Walking on a flat surface, b) Going up or down stairs, c) At night while in bed, d) Sitting or lying, e) Standing upright*. Response options: None; Mild; Moderate; Severe; Extreme. Each of the five items is scored from 0 to 4. These scores are added to a 0-20 sum score, with a higher score indicating worse pain

### Missing data

We will apply multiple imputation to impute missing WOMAC pain domain responses, although rows with missing NRS or without any WOMAC pain domain responses will be omitted from the dataset. We will use chained equations to generate the imputation-enriched dataset using predictive mean matching with the mice package in R (R Core Team, 2023).(10) The imputation model will include both demographic (baseline) and other outcome variables.

### Assumptions for harmonisation

We will evaluate the assumptions necessary for harmonizing the WOMAC pain domain and Numerical Rating Scale (NRS) scores. Below, we outline each assumption and describe how it will be assessed.

#### Conceptual equivalence

Since both the NRS and the WOMAC pain domain explicitly assess pain intensity in the operated joint, we believe that both instruments aim to measure the same conceptual construct, i.e. pain intensity in the operated joint.

#### Psychometric validity of the separate instruments

The validity of the WOMAC pain domain, including unidimensionality and reliability, have been tested numerous times and will be assessed specifically for the Danish version (see Study 1). Similarly, the NRS has shown high test-retest reliability,(26) while it is impossible to test the NRS for unidimensionality, being a single-item instrument.

#### Distribution of scores

We will assess whether the WOMAC pain domain and NRS scores appear normally distributed, or at least similarly distributed, and without severe skewness, kurtosis, or floor/ceiling effects.

We will use density plots and Q–Q plots to visualise the score distributions, and report the Shapiro–Wilk test results, although large samples tend to produce statistically significant results even for very small deviations.

#### Monotonicity

To examine whether the relationship between WOMAC pain domain and NRS scores is monotonic, we will create and inspect a scatterplot with a smooth curve, using locally weighted scatterplot smoothing.

#### Correlation

We will examine the correlation between the NRS scores and WOMAC pain domain sum scores with Pearson and Spearman statistics. High correlations suggest that the measures are closely related and may be suitable for harmonization.

#### Subgroup invariance

The measures above should exhibit measurement invariance across different subgroups, i.e., age, sex, BMI, and type of surgery, meaning that the assumptions and statistics mentioned are similar between key subgroups. For age, we will dichotomise at age 70 (i.e. age under 70 vs. age 70+). For BMI, we will dichotomise at BMI 30 kg/m^2^.

### Harmonization of NRS and WOMAC Pain Domain

We will perform equipercentile linking to create a conversion table from WOMAC pain domain sum scores to NRS scores and vice versa. In equipercentile linking, percentile ranks between the WOMAC and NRS scales are matched, ensuring that the distributions of the two measures are aligned. This method relies on the empirical cumulative distribution functions to determine percentile ranks, which are then matched. The function allows us to create a conversion table that maps each unique WOMAC sum score to its equipercentile-linked NRS value and vice versa. In addition to the conversion table, we will also produce a scatterplot of observed NRS scores against WOMAC sum scores, overlaid with the equipercentile linking curve

To evaluate the accuracy of the equipercentile linking, we will use the following metrics:

1. Root Mean Squared Error (RMSE) and Mean Absolute Error (MAE) to quantify the discrepancy between observed and linked NRS scores.
2. Spearman’s rank correlation to assess the monotonic relationship between observed and linked NRS scores. We use ranked correlation rather than pearson correlation, because we do not assume linearity between WOMAC pain domain sum and NRS scores.
3. Concordance correlation coefficients (CCCs) and intraclass correlation coefficients (ICC). The CCC evaluates the agreement between observed and linked scores by considering both precision and accuracy, while ICC assesses the consistency or absolute agreement between the two sets of scores.
4. Bland-Altman plots to visualize the agreement between observed and predicted scores, including the mean difference (bias) and 95% limits of agreement.
5. Residual plots to examine the distribution of residuals across the range of predicted scores.
6. Calibration Plot: A scatter plot of linked versus observed NRS scores, overlaid with a linear regression line and a 45-degree reference line. This allows visual assessment of how well the linked scores calibrate with the observed scores (with the reference line reflecting perfect calibration).

**Table 2:** Equipercentile conversion table from WOMAC pain domain to NRS scores, including number of respondents with each WOMAC pain domain score. Mock table:

| *WOMAC*<br>*pain*<br>*domain*<br>*score* | *n* | *Percentile* | *Linked*<br>*NRS*<br>*score* |
| --- | --- | --- | --- |
| 0 | 100 | 4 | 0 |
| 1 | 200 | 12 | 0 |
| 2 | 400 | 21 | 1 |
| 3 | ... | ... | ... |

## Tables and Figures

Table 1: A table with demographics, item missingness, and WOMAC pain domain sum score.

Table 3: Equipercentile conversion table from NRS score to WOMAC pain domain score, including number of respondents with each NRS score.

Figure 1: Flow of participant response data

Figure 2: A cross-walk plot – i.e. a scatterplot showing the distribution of WOMAC pain domain scores and NRS scores, overlayed by the equipercentile linking function.

## Knowledge dissemination

The results will be submitted for publication in a peer-reviewed journal. We will seek to make the reports freely available, either by open-access publication or through publication on a preprint server, e.g. www.medrxiv.org.

## Ethical Considerations

The project is listed on the Capitol Region of Denmark’s research listing (p-2025-19417), which by delegation from The Danish Data Protection Agency approves the handling of confidential data for research. According to Danish legislation, approval from the national ethics committee is not required for studies that do not collect biological samples or impose interventions (Appendix 1)

Only Jens Laigaard will have access to the data, which are stored, handled and analysed pseudonymised at a logged and encrypted drive. For studies using data from the Danish National Prescription Registry, data are uploaded to Statistics Denmark’s secure online research platform in order to link the data.(11) This remote-access environment can only be accessed with the corresponding author’s unique digital signature.

## Data availability statement

The data used in this study are available from their primary sources upon reasonable request. Please see the original publications for details.

## Appendix 1

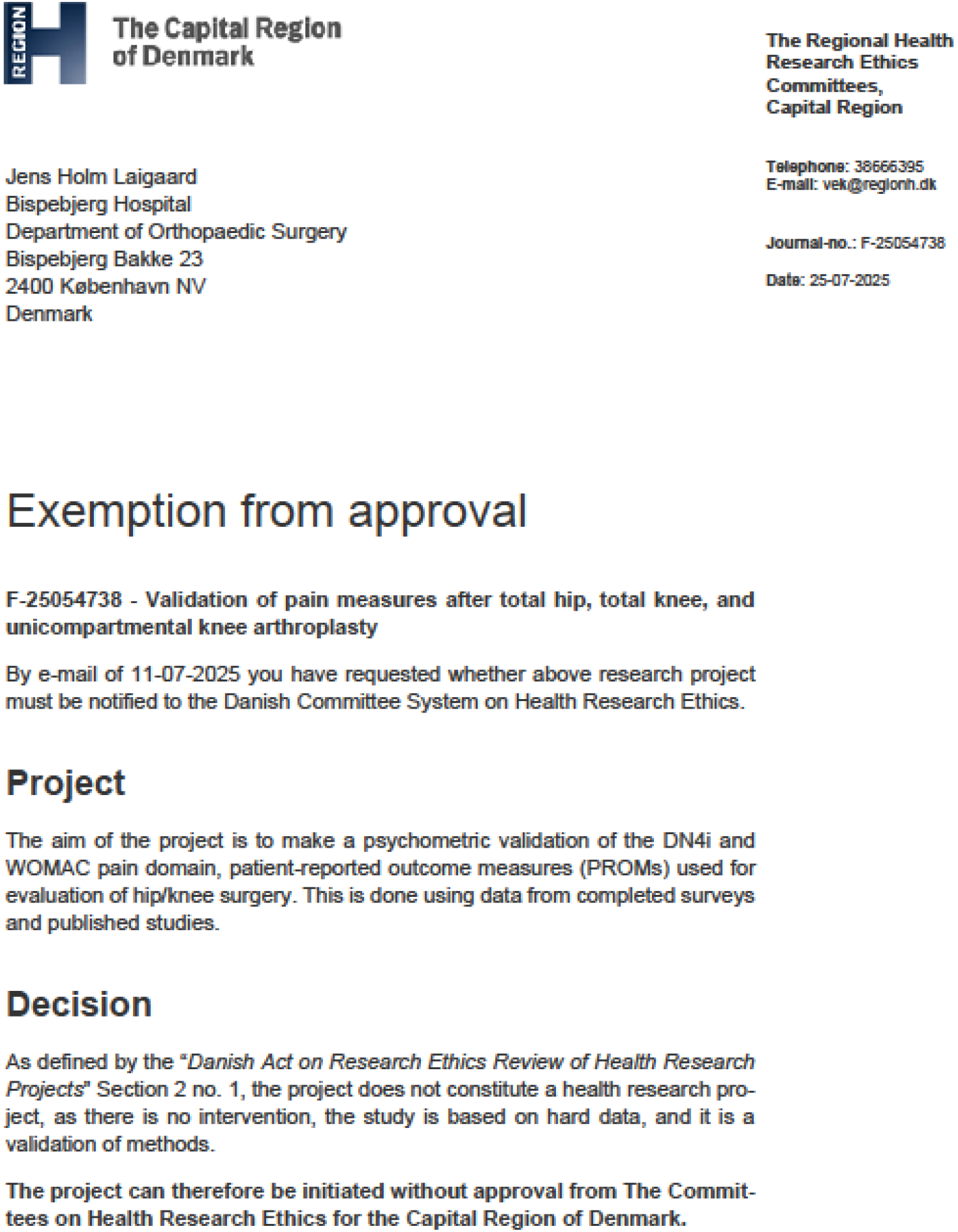

